# Global Trends of Seroprevalence and Universal Screening Policy for Chagas Disease in Donors: a systematic review and meta-analysis

**DOI:** 10.1101/2019.12.25.19015776

**Authors:** Jenny Yeon Hee Kim, Julia Ledien, Eliana Rodriguez-Monguí, Andy Dobson, María-Gloria Basáñez, Zulma M. Cucunubá

## Abstract

**Background:** Screening for *Trypanosoma cruzi* among blood and organ donors is essential to reduce Chagas disease transmission. The World Health Organization (WHO) has prioritised curtailing transmission in blood banks (BBs) and transplantation centres (TCs) by 50% by 2025 and 100% by 2030. This study aims to update the situation on *T. cruzi* screening strategies in BBs and TCs to evaluate the evolution of seroprevalence and the achievement of screening milestones globally.

**Methods:** We used published articles and government reports on seroprevalence data and screening policies in BBs and TCs across the world. We conducted meta-analyses of *T. cruzi* seroprevalence estimates by who region, endemicity status, and country, and used meta-regression to identify the covariates influencing the estimates. Publication bias and sensitivity analyses were also conducted.

**Results:** Based on 99 studies and reports and found a global pattern of increased universal screening policies (USPs) in BBs from 1990 to 2018. We found information for 50 countries, of which 44 (88%) have implemented USPs and 21 (42%) achieved 100% coverage by 2015. Out of the 21 Chagas-disease endemic countries, 20 are in advanced USPS stages, and 18 achieved 100% coverage by 2015. Latin America (LA) was the first region to start USPS since the 1990s and 19 countries are in advanced stages of implementation and by 2015 there is evidence of 100% coverage in 15 LA countries. In the Caribbean Region, USPs are still in early implementation stages and by 2015 only five out of 24 countries have achieved 100% coverage. Outside Latin America and the Caribbean, there are USPs only in the USA, which initiated in 2007 and with 100% coverage in 2016. In Europe, there are no USPs, but some countries have implemented selective screening of at-risk donors in the UK, Spain, France and Switzerland. Whereas Sweden and Italy have implemented a deferral system. For TCs, national guidelines have been produced in some European countries since the 2000s; in the USA, USPs started since 2002, but 100% coverage is yet to be achieved. There is a global decrease in *T. cruzi* seroprevalence among blood donors from the 1970s to 2010s, particularly in endemic countries, where the *T. cruzi* pooled seroprevalence decreased from 2.42% (95% CI 0.75%-7.53%) in the 1970s to 0.38% (95% CI 0.30%-0.60%) in the 2010s. Seroprevalence in non-endemic countries has remained relatively stable between 1990s and 2010s around 0.01% (95% CI 0.01%-0.03%). Country and decade were identified as the two major predictors of seroprevalence in BBs. Data on TCs was scarce.

**Interpretation:** Despite global progress in *T. cruzi* screening policies, both USPs and 100% coverage are yet to be achieved. Seroprevalence in BBs have decreased in endemic countries, likely due to a combination of vector control, increased USPs and voluntary donation, and improved diagnosis. To achieve the proposed WHO goals by 2025 and 2030, USPs in TCs must become available in all endemic countries. In BBs, USPs should be a priority in the Caribbean region as well as non-endemic countries where migration from endemic countries is important.

## Introduction

Chagas disease (CD), also known as American trypanosomiasis, is a disease caused by *Trypanosoma cruzi*, a parasitic protozoan mainly transmitted by triatomine vectors (“kissing bugs”). It can also be transmitted via food contamination, blood transfusion, organ transplantation, or congenitally ^1^. During the chronic phase of CD, 20-30 % of those infected may develop life-threatening complications, including cardiac and neurological disorders that can lead to death ^2–4^

Approximately 8 million people are infected with *T. cruzi* worldwide ^3^. The Pan-American Health Organization (PAHO) estimates that currently about 6 million people are infected with *T. cruzi* in Latin America, with 28,000 new cases and 12,000 deaths occurring per year^5^. Due to recent rising international travelling and migration, the epidemiological profile of CD in non-endemic countries is changing. In 2015, it was estimated that there were 42,000 cases in Spain, 1,712 in France, and 1,324 in the United Kingdom ^6^. Currently, the CDC indicates that there are >300,000 cases in the USA^7^.

In the 1990s, the governments of the six CD-endemic Southern Cone countries (Argentina, Bolivia, Brazil, Chile, Paraguay and Uruguay) where Chagas disease is endemic initiated the integrated efforts to eliminate *T. cruzi* vectors via insecticide spraying and housing improvement (known as the Southern Cone Initiative). As a result, vector infestation was significantly reduced in these countries ^8,9^. Following the initial success of vector-control programmes, blood transfusion from infected untreated donors emerged as the second most common transmission route^10–12^. In response, governments mandated the serological screening for *T. cruzi* of blood donations or donors after the Southern Cone Initiative and, by 2005, twelve of the CD-endemic Latin American countries had achieved 100% screening coverage. With the increasing cases of transfusion-transmitted infections, governments of non-endemic countries have recognized the importance of implementing strategies for controlling the transmissions ^13,14^; the UK, France, Spain, and the USA have initiated screening of blood banks (BBs) since the early 2000s ^14^.

Currently, the World Health Organization (WHO) goals for CD include achieving complete interruption of *T. cruzi* transmission in BBs and transplantation centres (TCs) by 2030 ^15^. Despite the availability of national guidelines for *T. cruzi* screening in BBs and TCs, it remains unknown whether such guidelines have been implemented and (or) to which extent recommendations have been followed, making it difficult to evaluate progress towards the WHO 2030 goals. Therefore, we conducted a systematic review and meta-analysis of screening policies and seroprevalence data in BBs and TCs globally to quantify and compare progress in the implementation of such policies.

### Methods

### Search Strategy and Selection Criteria

We conducted a systematic review on screening policies and *T. cruzi* seroprevalence data and subsequent meta-analyses of the obtained data from in BBs and TCs, with no restrictions on language, country, time, age or gender. The PRISMA (Preferred Reporting Items for Systematic Reviews and Meta-Analyses) guidelines and checklist were used (appendix pp 2).

We systematically searched databases including Medline (a subset of PubMed), Web of Knowledge, Literatura Latino-Americana e do Caribe em Ciencias da Saúde (LILACS), Google Scholar and Excerpta Medica (EMBASE) databases. We included studies describing country’s implementation of *T. cruzi*-screening strategies, presence of *T. cruzi* seroprevalence data, and any relevant model-derived estimates of *T. cruzi* seroprevalence among blood donors, organ donors and recipients. Inclusion and exclusion criteria are available in appendix pp 2. Our search algorithm combined the search terms: 1) blood bank, 2) blood bank screening 3) blood donor 4) transfusion, 5) transplant, 6) organ transplantation, 7) Chagas disease, 8) *Trypanosoma cruzi* (appendix pp 4-5).

We imported the selected citations into the online software Covidence ^16^. All titles and abstracts were reviewed by two independent investigators and conflicts were resolved by a third person. Abstracts were screened in Spanish, Portuguese, English, and French. Full-text screening was also conducted in the same manner.

### Data extraction

Each paper selected for full-text screening was reviewed carefully and the relevant quantitative and qualitative information was extracted regarding screening policy and seroprevalence. Extracted data included the information on year of screening policy implementation, type of screening policy (universal screening, selective screening, permanent deferral), country of policy implementation, and any changes on the existing screening policies. For seroprevalence, relevant extracted data included year of publication, year of study, country, city/region, CD endemicity status (endemic/non-endemic), pre-test risk of infection, name of the BB/TC, case definition, pre-screening criteria, sampling method, age range, diagnostic technique, number of conducted tests, and number of positives. When point estimates were not provided in the papers, these were calculated using the extracted data. We classified the screening policies into four categories: a) deferral (i.e. deferred from blood donation when person may think was at risk), b) selective screening of at-risk populations, universal policy screening (USP) and d) not clear (Table S3, appendix pp 6). USPS was classified according to achieved coverage.

Data quality assessment was conducted based on a checklist of criteria used by Ding et al ^17^. We evaluated the research question, sampling method, study period, diagnostic method, and any potential bias within the selected studies. For each study, the criteria were considered to allocate a score to classify the data as of high, moderate, or low-quality (Table S4, appendix pp 8).

### Data analysis

Data analysis consisted of three components: meta-analysis, publication bias assessment, and meta-regression. Taking into account that each study may differ in geographic location, pre-test infection risk, and CD endemicity, a random-effects model was used. We estimated the overall and decade-specific seroprevalence. Differences in the seroprevalence between decades was considered significant if test for group differences X^2^ had a p value < 0.05. We report *I*^*2*^ statistic as the proportion of observed variance among the study results due to heterogeneity ^18^ and tau-squared (*τ*^*2*^) to indicate the extent of between-study variance ^18^.

In addition to an overall meta-analysis, we also conducted meta-analyses by the WHO sub-regions for which we could find the data within selected papers (Europe, North America, Meso America, Caribbean and South America), endemicity, and country. Within the meta-analyses by CD endemicity, non-endemic countries include North American and European countries, whereas endemic countries include Meso American and South American countries. Meta-analyses by country include the countries with at least five reports on seroprevalence at blood banks. We conducted the statistical analyses of collected data using R v.3.5.3 ^19^.

Publication bias was assessed using funnel plots and Egger’s regression asymmetry test ^20^. We also used a trim and fill technique to identify and correct the asymmetry in funnel plots by firstly removing the smaller studies causing the funnel plot asymmetry, secondly estimating the true centre of the funnel, and finally filling the missing studies around the centre to calculate the adjusted study effects.

We conducted a meta-regression to identify the covariates that may influence the pooled seroprevalence values and produce a model that best predicted the variability of effect sizes. We selected the model that maximised maximum likelihood, minimised the Akaike Information Criterion (IC), and explained the highest percentage of the variance of the dependent variable (R^2^).

### Findings

The systematic literature search identified a total 2,830 studies, which were identified according to the selection criteria. Ninety-nine studies were selected for data extraction, of which sixty-two studies were based on seroprevalence data from BBs, nineteen studies from TCs, and seven studies from reports on transfusion-transmitted infections. Eleven references were selected for review of screening policies across countries (Figure 1). Further details on selected studies are available in supplementary materials (Table S6, appendix pp 11). The references for all studies used in the analyses are also listed in supplementary materials (appendix pp 19).

**Figure 1.**
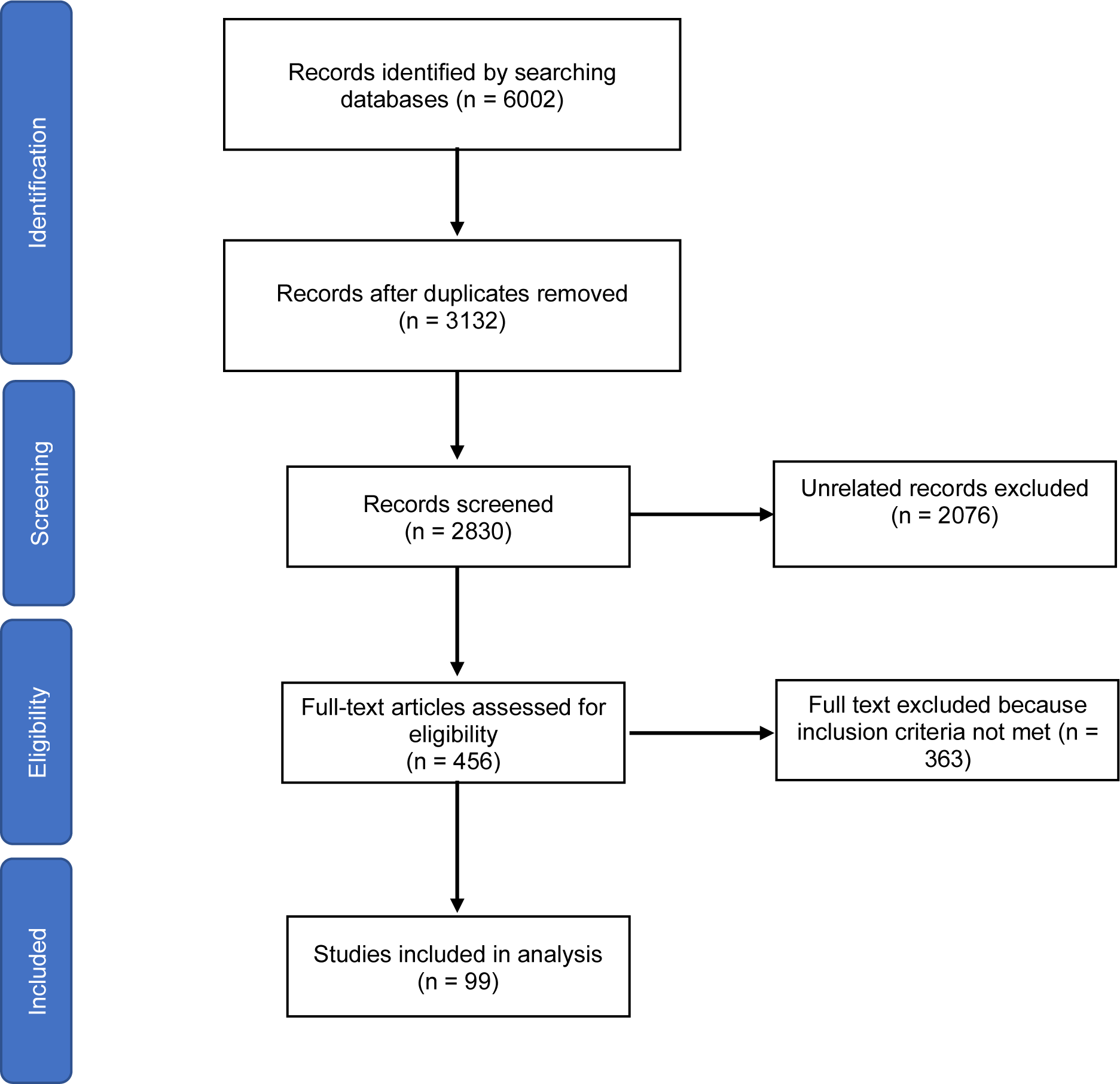
Flow diagram describing the process of selecting the relevant studies for meta-analysis

From the data-quality assessment, of 99 selected references, 1 showed low quality, 32 showed moderate quality, and 63 showed high quality. See supplementary materials (Table S5, appendix pp 8) for the full quality assessment. A summary of the total number of reports on seroprevalence and individuals included per country according to WHO region, sub-region ^21^, and country, is presented in Table 1.

**Table 1.** List of countries and reports included in the meta-analysis of *T. cruzi* seroprevalence in blood banks

| WHO region | Sub-region | Country | Number of reports | Year of first study | Total | Positives |
| --- | --- | --- | --- | --- | --- | --- |
| Americas | North America | Canada | 3 | 1997 | 90,208 | 14 |
|  |  | United States | 15 | 1993 | 37,448,510 | 1,767 |
|  | Meso America | Mexico | 22 | 1993 | 11,487,085 | 2,116 |
|  |  | Costa Rica | 12 | 1988 | 579,518 | - |
|  |  | El Salvador | 12 | 1993 | 1,634,798 | 21,693 |
|  |  | Guatemala | 7 | 1993 | 673,710 | - |
|  |  | Honduras | 12 | 1993 | 520,845 | - |
|  |  | Nicaragua | 11 | 1993 | 627,405 | - |
|  |  | Panama | 10 | 1994 | 456,366 | - |
|  | South America | Argentina | 22 | 1987 | 7,244,988 | 47,599 |
|  |  | Bolivia | 15 | 1988 | 549,645 | 321 |
|  |  | Brazil | 33 | 1985 | 18,964,142 | 17,438 |
|  |  | Chile | 23 | 1982 | 1,884,824 | 888 |
|  |  | Colombia | 13 | 1993 | 5,586,353 | 637 |
|  |  | Ecuador | 13 | 1992 | 1,653,875 | 55 |
|  |  | Paraguay | 12 | 1993 | 612,888 | - |
|  |  | Peru | 12 | 1993 | 1,329,171 | 119 |
|  |  | Uruguay | 10 | 1994 | 1,032,212 | - |
|  |  | Venezuela | 37 | 1981 | 3,413,414 | 1,375 |
| Caribbean Region | Caribbean Region | Belize | 3 | 1977 | 10,855 | 5 |
|  |  | Bermuda | 2 | 2014 | 3,278 | - |
|  |  | Guadeloupe | 1 | 2015 | 7,891 | - |
|  |  | Guyana | 2 | 2007 | 11,702 | 7 |
|  |  | Martinique | 1 | 2015 | 11,217 | - |
|  |  | Suriname | 2 | 2014 | 20,811 | - |
| Europe | Western Europe | France | 1 | 2007 | 30,837 | 3 |
|  |  | Switzerland | 2 | 2011 | 7,707 | 3 |
|  | Southern Europe | Spain | 2 | 2005 | 1,933 | 14 |
|  |  | Italy | 2 | 2008 | 156 | 5 |
|  | Northern Europe | United Kingdom | 1 | 1998 | 38,585 | 3 |
| Western Pacific | Southeast Asia | Japan | 1 | 2004 | 18,076 | 3 |

**Table 2.** List of findings from individual meta-analyses on the countries with at least 5 observations reported

| Sub-region | Country | Decade | Reports (n) | Estimated Pooled Prevalence% (95% CI) | pval (X<sub>2</sub>) | I<sub>2</sub> (%) | τ<sub>2</sub> | pval (Q) |
| --- | --- | --- | --- | --- | --- | --- | --- | --- |
| North America | United States | **Total** | **12** | **0.01 (0.01-0.03)** | **0.06** | **100** | **2.38** | **<0.01** |
|  |  | 1990s | 4 | 0.06 (0.01-0.37) |  |  |  |  |
|  |  | 2000s | 5 | 0.01 (0.00-0.01) |  |  |  |  |
|  |  | 2010s | 3 | 0.01 (0.01-0.02) |  |  |  |  |
| Meso America | Costa Rica | **Total** | **10** | **0.30 (0.14-0.65)** | **0.03** | **100** | **1.48** | **<0.001** |
|  |  | 1990s | 4 | 0.78 (0.23-2.57) |  |  |  |  |
|  |  | 2010s | 6 | 0.17 (0.10-0.30) |  |  |  |  |
|  | El Salvador | **Total** | **11** | **2.14 (1.88-2.44)** | **<0.01** | **99** | **0.05** | **<0.01** |
|  |  | 1990s | 4 | 2.17 (2.02-2.34) |  |  |  |  |
|  |  | 2000s | 1 | 2.45 (2.42-2.48) |  |  |  |  |
|  |  | 2010s | 6 | 2.08 (1.66-2.680) |  |  |  |  |
|  | Guatemala | **2010s** | **6** | **1.05 (0.96-1.15)** | **NA** | **94** | **0.01** | **<0.01** |
|  | Honduras | **Total** | **10** | **1.23 (1.05-1.44)** | **0.01** | **98** | **0.07** | **<0.01** |
|  |  | 1990s | 4 | 1.48 (1.28-1.70) |  |  |  |  |
|  |  | 2010s | 6 | 1.09 (0.90-1.32) |  |  |  |  |
|  | Mexico | **Total** | **15** | **0.50 (0.31-0.79)** | **0.41** | **100** | **0.84** | **<0.001** |
|  |  | 1990s | 2 | 0.52 (0.32-0.84) |  |  |  |  |
|  |  | 2000s | 4 | 0.84 (0.21-3.30) |  |  |  |  |
|  |  | 2010s | 9 | 0.39 (0.27-0.55) |  |  |  |  |
|  | Nicaragua | **Total** | **10** | **0.34 (0.29-0.41)** | **<0.01** | **94** | **0.07** | **<0.01** |
|  |  | 1990s | 4 | 0.44 (0.40-0.50) |  |  |  |  |
|  |  | 2010s | 6 | 0.29 (0.25-0.34) |  |  |  |  |
|  | Panama | **Total** | **9** | **0.50 (0.34-0.73)** | **0.86** | **99** | **0.34** | **<0.01** |
|  |  | 1990s | 3 | 0.53 (0.17-1.63) |  |  |  |  |
|  |  | 2010s | 6 | 0.48 (0.41-0.56) |  |  |  |  |
| South America | Argentina | **Total** | **16** | **3.44 (2.26-5.21)** | **<0.001** | **100** | **0.78** | **<0.001** |
|  |  | 1980s | 4 | 6.23 (3.73-10.20) |  |  |  |  |
|  |  | 1990s | 1 | 3.23 (2.06-5.02) |  |  |  |  |
|  |  | 2000s | 2 | 3.75 (0.27-36.34) |  |  |  |  |
|  |  | 2010s | 6 | 2.33 (1.92-2.83) |  |  |  |  |
|  | Bolivia | 2010s | 6 | 2.77 (2.34-3.26) | NA | 99 | 0.05 | <0.01 |
|  | Brazil | **Total** | **31** | **0.42 (0.25-0.70)** | **<0.001** | **100** | **2.13** | **<0.001** |
|  |  | 1980 | 5 | 3.39 (2.60-4.41) |  |  |  |  |
|  |  | 1990s | 8 | 0.79 (0.48-1.29) |  |  |  |  |
|  |  | 2000s | 7 | 0.20 (0.08-0.52) |  |  |  |  |
|  |  | 2010s | 11 | 0.17 (0.09-0.29) |  |  |  |  |
|  | Chile | **Total** | **10** | **0.86 (0.39-1.92)** | **<0.001** | **100** | **2.72** | **<0.001** |
|  |  | 1980 | 7 | 4.21 (2.19-7.94) |  |  |  |  |
|  |  | 1990s | 2 | 0.98 (0.96-1.01) |  |  |  |  |
|  |  | 2000s | 1 | 0.40 (0.34-0.48) |  |  |  |  |
|  |  | 2010s | 6 | 0.15 (0.13-0.17) |  |  |  |  |
|  | Colombia | **Total** | **10** | **0.71 (0.47-1.08)** | **<0.01** | **100** | **0.45** | **<0.001** |
|  |  | 1990s | 3 | 1.55 (1.02-2.34) |  |  |  |  |
|  |  | 2010s | 7 | 0.51 (0.37-0.72) |  |  |  |  |
|  | Peru | **Total** | **9** | **0.50 (0.21-1.18)** | **<0.01** | **100** | **1.75** | **<0.01** |
|  |  | 1970 | 1 | 5.71 (4.37-7.44) |  |  |  |  |
|  |  | 1990s | 2 | 0.08 (0.02-0.30) |  |  |  |  |
|  |  | 2010s | 6 | 0.61 (0.47-0.79) |  |  |  |  |
|  | Uruguay | **Total** | **10** | **0.36 (0.27-0.49)** | **<0.01** | **99** | **0.22** | **<0.01** |
|  |  | 1990 | 4 | 0.61 (0.59-0.64) |  |  |  |  |
|  |  | 2010s | 6 | 0.25 (0.21-0.31) |  |  |  |  |
|  | Venezuela | **Total** | **13** | **0.90 (0.47-1.71)** | **<0.001** | **100** | **1.43** | **<0.001** |
|  |  | 1970 | 1 | 4.00 (3.95-4.05) |  |  |  |  |
|  |  | 1980 | 2 | 6.21 (1.32-24.71) |  |  |  |  |
|  |  | 1990 | 4 | 0.91 (0.73-1.13) |  |  |  |  |
|  |  | 2000s | 1 | 0.74 (0.67-0.82) |  |  |  |  |
|  |  | 2010s | 5 | 0.31 (0.29-0.34) |  |  |  |  |
| **A country estimate is given only if the number of studies is >= 5**<br>**pval(X<sub>2</sub>):** p value from test for decade differences<br>**I<sub>2</sub>:** measure of the proportion of observed variance among the study results due to heterogeneity<br>**τ<sub>2</sub>:** value reflecting the extent of variation among the study effects<br>**pval(Q):** p value for heterogeneity |  |  |  |  |  |  |  |  |

### Universal Screening Policies in Blood Banks and Transplantation Centres

From the selected eleven papers and reports which described the evolution of *T. cruzi* screening policies, we found information for 50 countries out of which USPs has been implemented in 44. The evolution *T. cruzi* USPs implementation over time shows a clear pattern of increase since 1990 to 2015 (Figure 2). However, out of 50 only 21 countries have achieved 100% coverage by 2015. Importantly, all 21 Chagas disease endemic countries are in advanced states of USPs implementation. By 2015, 18 of them have reached 100% coverage.

**Figure 2.**
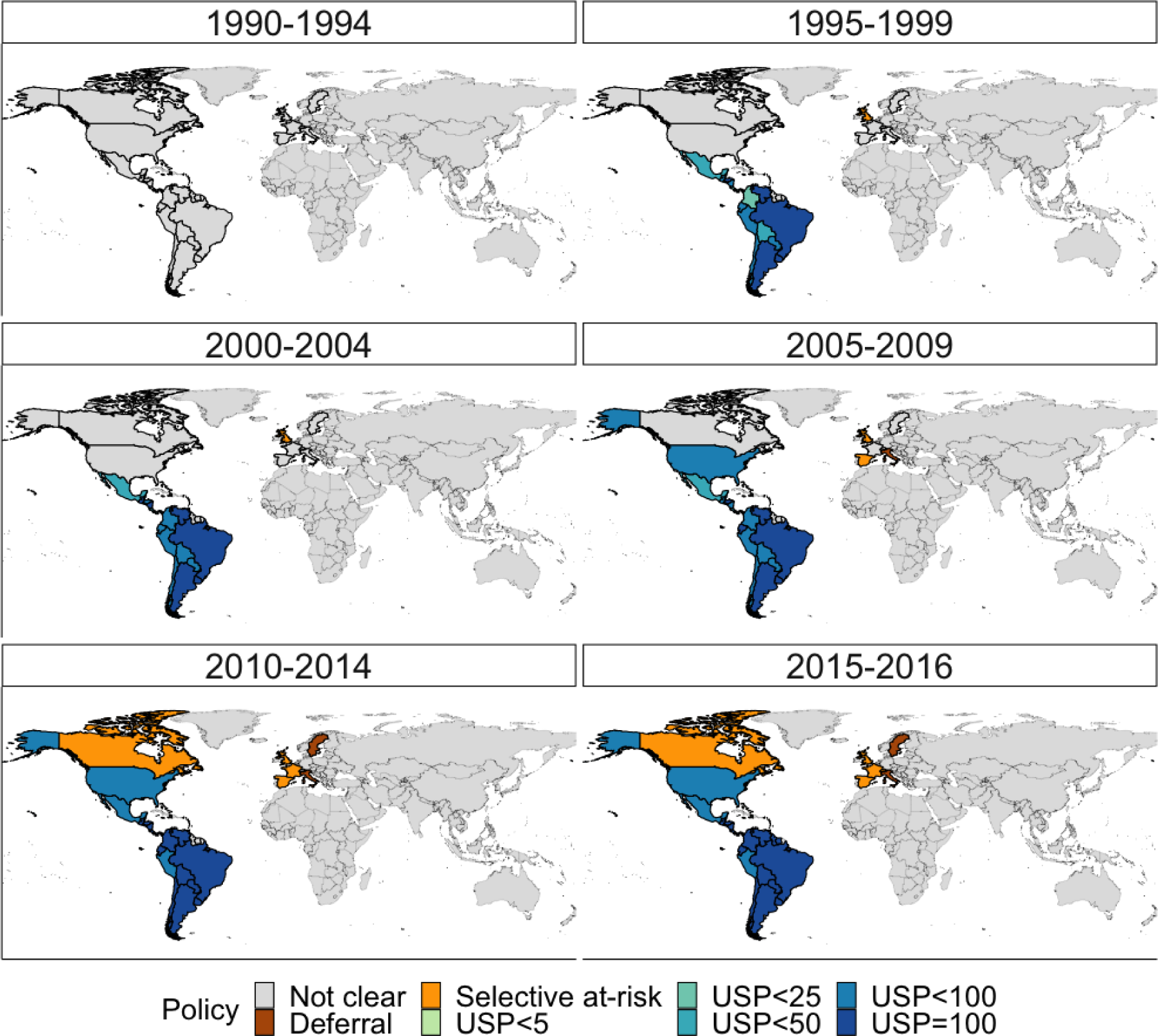
Global changes in blood donor screening policy for T. cruzi from 1990 to 2016, towards universal screening policy (USP)

In North America, the USA initiated the USPs in 2007 ^21^, reaching 100% of coverage in all BBs by 2016 ^22^. Canada began screening the at-risk donors in 2010, using the questionnaires assessing the risk factors^11^.

Latin America was the first region to start USPs, of which all countries have initiated since the 1990s ^11^, but each country has reached 100% of coverage in different years ^23–25^. By the year 2015, we observed the achievement of 100% coverage in universal screening in most of the Latin American countries, except for Mexico and Peru.

In the Caribbean region, most countries have initiated USPs in the 2010s, but only five out of 24 countries reached 100% of coverage ^25^. By 2015, there is evidence of the achievement of 100% of coverage in universal screening in Belize, Guyana, Suriname, Martinique, and Trinidad and Tobago.

There currently is no USPs in Europe, but some countries have implemented the selective screening of at-risk donors in BBs using the risk factor questionnaires in the UK, since 1995; Spain, since 2005; France, since 2009, and Switzerland, since 2013. In Sweden, systematic screening of blood donors has not been implemented; however, all donors who have lived in endemic countries for more than 5 years are permanently deferred from blood donations ^11,14^.

Italy, Spain, and the UK have created the national guidelines on controlling the CD transmissions by organ transplantations via routine *T. cruzi* screening starting from the year 2012, 2004, and 2011, respectively ^14^. Since 2002, organ procurement organizations in the USA have initiated partial coverage of universal screening for all tissue donors^26^. See supplementary materials for the map (Figure S1, appendix pp 19).

### Seroprevalence in blood banks

We were only able to find seroprevalence data on European, North American, and Latin American and Caribbean regions. The overall meta-analysis, conducted with all studies, showed a decrease in seroprevalence from 2.42% (95% CI 0.75%-7.53%) in the 1970s to 0.38% (95%CI 0.30%-0.49%) in the 2010s. During this period, differences between decades were statistically significant (p < 0.001). The pooled seroprevalence over a five-decade period (from 1970s to 2010s) was 0.48% (95% CI 0.39%-0.60%) (*I*^2^= 100%, τ^2^=3.05, p < 0.001) (Figure S2 appendix pp 20)

[3.20; 6.42]

In endemic countries there is a significant (p < 0.01). decrease in seroprevalence from 4.55% (95% CI 3.20%-6.42%) in the 1980s to 0.49% (95% CI 0.40%-0.60%) in the 2010s (Figure 3A, and Figure S3 appendix pp 20). By contrast, when analysing only non-endemic countries, including Canada, France, Japan, Spain, Switzerland, and the USA, there is a non-significant decrease (p=0.33) in seroprevalence from 0.04% (95% CI 0.01%-0.26%) in the 1990s to 0.01% (95% CI 0.02%-0.03%) in the 2010s (Figure 3B, Figure S4, appendix pp 21).

**Figure 3.**
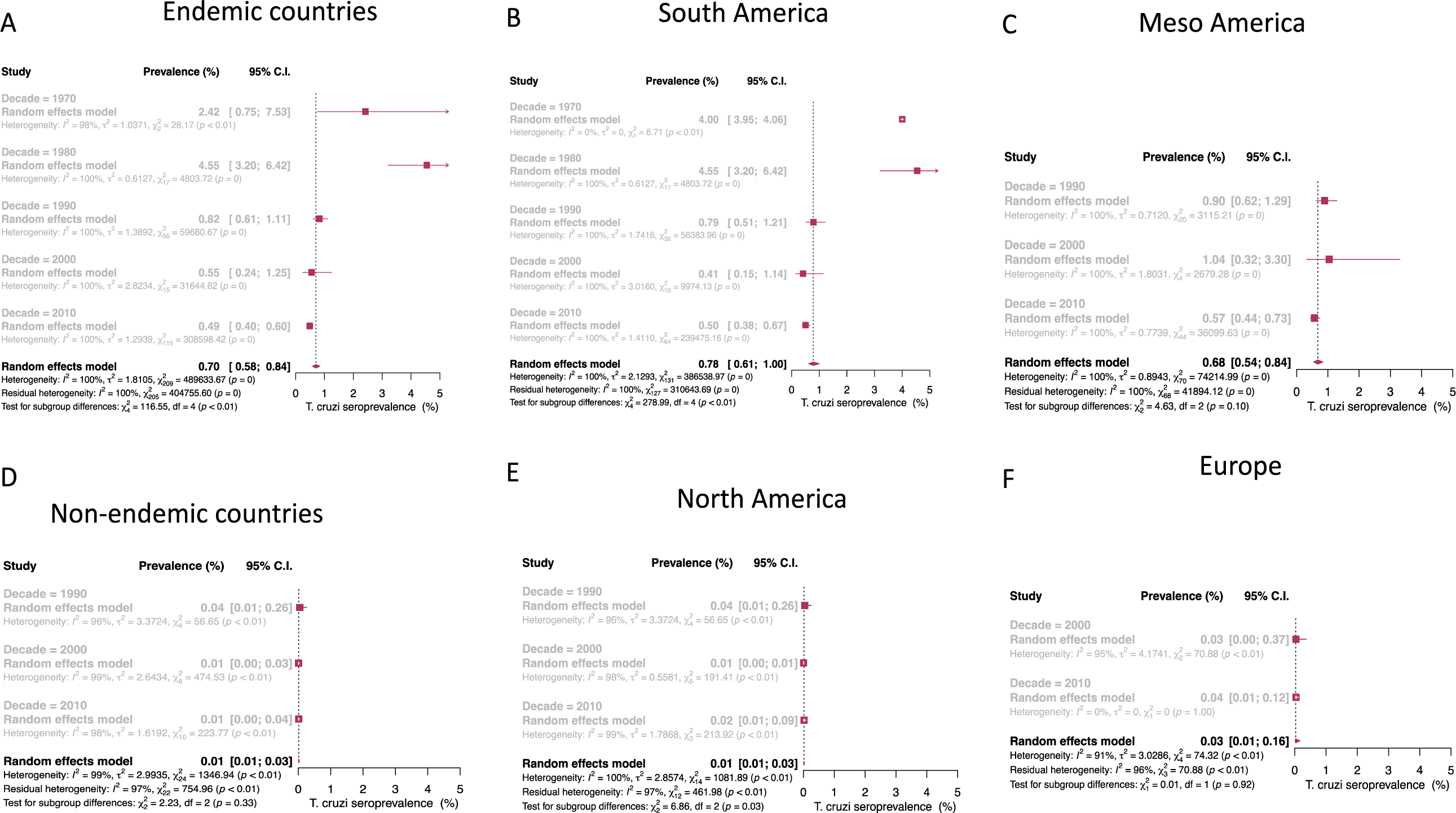
Forest plot of the meta-analysis of *Trypanosoma cruzi* seroprevalence in A) Endemic countries, B) South America, C) Meso America, D) Non-endemic countries, E) North America and F Europe. Red squares represent central estimate.

In the South America region, including Argentina, Bolivia, Brazil, Chile, Colombia, Ecuador, Paraguay, Peru, Uruguay, and Venezuela, showed a statistically significant (p < 0.01) decrease in seroprevalence from 4.00% (95% CI 3.95%-4.06%) in the 1970s to 0.50% (95% CI 0.38%-0.67%) in the 2010s (Figure 3C and Figure S5 appendix pp 21).

The Meso America region, included Mexico, Costa Rica, El Salvador, Guatemala, Honduras, Nicaragua, and Panama, showed a statistically non-significant decrease in seroprevalence from 0.90 % (95% CI 0.62%-1.29%) in the 1990s to 0.57% (95% CI 0.44%-0.73%) in the 2010s (Figure 3D and Figure S6 appendix pp 22).

In the Caribbean region, including Belize, Guyana, Bermuda, Guadeloupe and Martinique there was only one report available in the 1970s and 10 reports for 2010s. The current seroprevalence for the region was estimated at 0.01% (95% CI 0.004% - 0.12%) (*I*^2^ = 96% %, τ2 = 6.4, p = 0.04) (Figure S7 appendix pp 20).

The North America region, including Canada and the USA, showed a statistically significant (p = 0.03) decrease in seroprevalence (of 50%) from 0.04% (95% CI 0.01%-0.26%) in the 1990s to 0.02% (95% CI 0.01%-0.09%) in the 2010s (Figure 3E and Figure S8 appendix pp 23).

The Europe region, including France, Spain, Switzerland, and the UK, showed a similar seroprevalence of 0.03% (95% CI 0.00%-0.37%) in the 2000s to 0.04% (95% CI 0.01%-0.12%) in the 2010s. The pooled seroprevalence over a two-decade period (2000s and 2010s) was 0.03% (95% CI 0.01%-0.16%) (*I*^2^ = 91%, τ^2^ = 3.03, p < 0.01) (Figure 3F and Figure S9 appendix pp 23).

With the 15 countries from where at least five reports were available, we conducted individual country meta-analysis. Among them, in endemic regions, the highest contemporary (2010s) sero-prevalence was found in Argentina 2.77% (2.34%-3.26%), followed by Bolivia at 2.33 % (1.92%-2.83%) and El Salvador at 2.08 % (1.66-2.68). Whereas in non-endemic countries, estimates where only possible in one country, the US with seroprevalence at 0.01% (0.01%-0.02%). Individual country results are presented in Table 3.

**Table 3.**
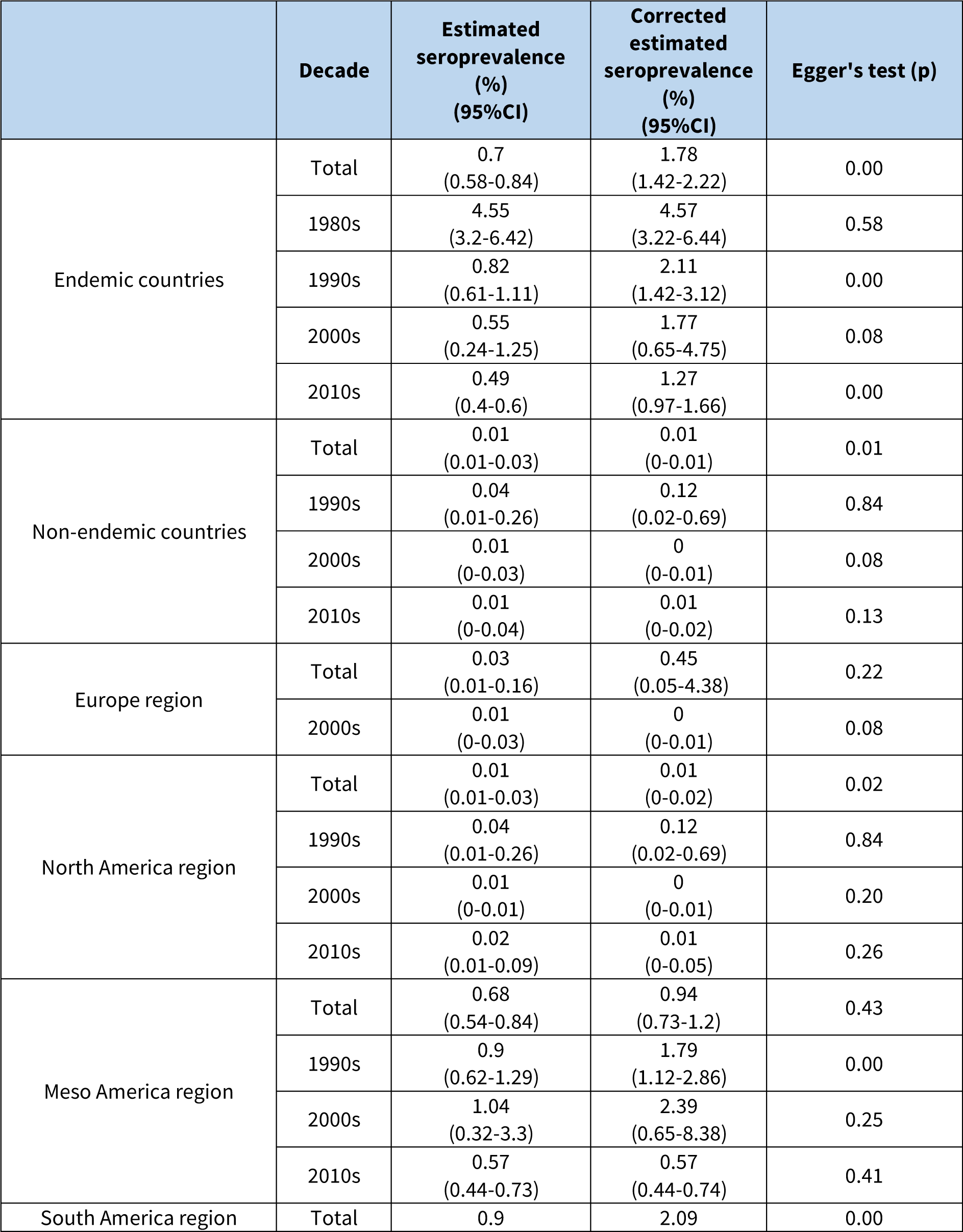
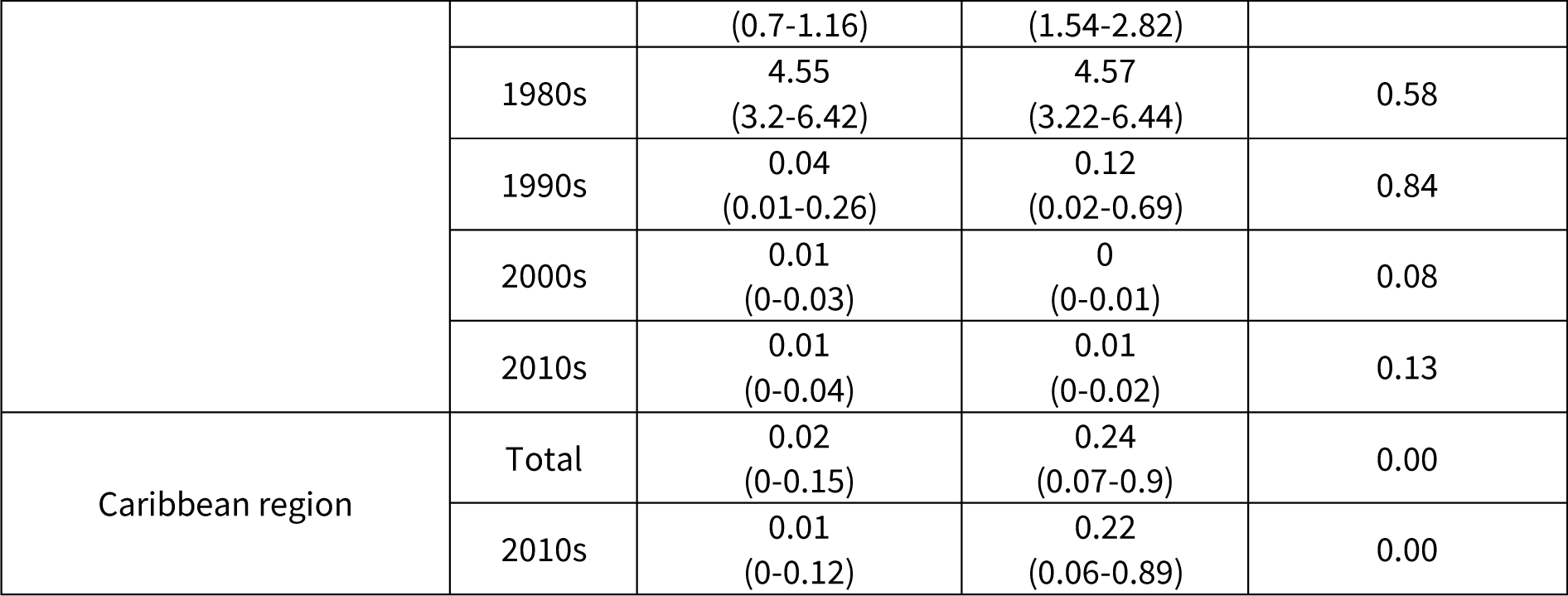
List of results from Trim and Fill technique, and Egger’s test

Analysis of publication bias indicated that such bias has a stronger influence when the time component is not considered and is less evident when the analysis is separate into decades (Table 3). See supplementary materials (Figure S10 appendix pp 24) for funnel plots.

From five meta-regression models the combination of variables ‘decade’ and ‘country’ best predicted the pooled *T. cruzi* seroprevalence (Table S7 and S8 appendix pp17).

## Discussion

With the aim to evaluate the degree of achievement of CD screening goals in BBs and TCs, our study has identified and extracted the relevant data to provide overall trends in *T. cruzi* seroprevalence and implementation of screening policies in both endemic and non-endemic countries over the past five decades. Despite the evident progress in the number of countries that have initiated or consolidated USPs, the essential implementations are yet to achieve their universal coverage. Out of 50 countries analysed, USPs have been implemented in 44 countries but only 21 have achieved 100% coverage.

Our study indicates a gradual and positive trend towards implementation of screening with increasing coverage across decades in North American and Latin American countries, as well as a few a few Caribbean and European countries. By 2015, most Latin American countries and the USA reached 100% coverage of universal BB screening for *T. cruzi* whereas the remaining countries established other screening or control measures. Among the policies regarding the control of transmissions by organ transplantations, we are only able to report the implementation of national guidelines in Italy, Spain, and the UK. However, due to paucity of data it is not possible to learn to which extent these recommendations are being followed by the health authorities of these countries.

From the overall meta-analysis, we estimated a global pooled seroprevalence of 0.48% (95% CI 0.39%-0.60%), but with markedly significant differences between the geographical regions and over five decades studied. Testing for differences between decades within the overall meta-analysis indicated a statistically significant difference in seroprevalence (p < 0.001). Such observation was consistent throughout the majority of country-level meta-analyses, which showed a general decline in the *T. cruzi* seroprevalence from the 1970s to 2010s. In particular, dramatic changes between and after the year 2000 imply possibly the impact of the interventions implemented by the Southern Cone Initiatives—we detected the greatest difference in the meta-analysis on endemic countries, where the *T. cruzi* pooled seroprevalence decreased from 4.52% (95% CI 2.96%-6.85%) in the 1980s to 0.70% (95% CI 0.59%-0.84%) in the 2010s.

Nevertheless, we hypothesise four possibilities to explain the estimated changes in the seroprevalence rates, the high heterogeneity and the significant decade effects. Firstly, the late 20^th^ and early 21^st^ centuries were marked by the regional initiatives to prevent CD transmission in Latin America, consisting in vector control programmes that markedly reduced domestic infestation indices, followed by legislation on *T. cruzi* universal screening in BBs, both impacting *T. cruzi* transmission^8,9^. Second, screening policies in blood banks may have first started in areas with the highest prevalence and as universal coverage is achieved, also lower endemic areas become included and so diluting the seroprevalence. Third, an increase in a number of voluntary non-remunerated blood donors (VNRBDs) could have contributed to improving blood safety. Since 2012, PAHO have reported an increase in the proportion of blood donations from voluntary and also repetitive donors in the Americas and also repetitive donation ^23–25^. Thus, low pooled seroprevalence estimates in the decades 2000s and 2010s across countries may also be due to the promotion of blood donations from the VNRBDs. Lastly, combinations of serological assays to test IgG against *T. cruzi* over the last five decades had varying degrees of sensitivity and specificity, with a trend for their diagnostic performance to improve over time. For example, the low specificity from *T. cruzi* crude antigens may have contributed to a higher rate of false positivity in CD diagnosis ^27–29^. Starting from the late 1980s, however, the quality control programmes for laboratory performances of serological tests encouraged the use of purified and recombinant antigens ^29,30^. Variations in types and quality of diagnostic assays and their qualities in different periods may, therefore, may also explain some of the reductions in the pooled seroprevalence estimates over time.

The change in seroprevalence in endemic countries shows also a very important step forward in screening policies in endemic countries where the seroprevalence shows an evident decrease and so a separation from the true population seroprevalence. This means the policies for voluntary and repetitive donations are having an effect on the risk of Chagas transmission via donations. Interestingly, this also means we cannot estimate Chagas population prevalence from screening in BBs, as we would be underestimating the true prevalence. Nevertheless, we could say that the true Chagas seroprevalence in these countries is at least as low as estimated in blood banks.

Although the study identified nearly a hundred articles for data extraction, there still remain the limitations on data availability, and particularly for TCs. Additionally, other variables, such as age, gender, types of blood donor, or sensitivity and specificity of diagnostic assays were not investigated due to this type of information missing in some of the studies. Notwithstanding these limitations, our study has several strengths. To our knowledge this is the first systematic review and meta-analysis conducted to answer the question on the progress of blood bank screening policies for Chagas disease worldwide. We also made efforts to minimize study biases with the meta-analyses by different variables, producing the results at both regional and national levels that were consistent with the chosen meta-regression model.

This study provides an in-depth view of the global status of screening policies as well as of *T. cruzi* seroprevalence in relation to BBs. Despite global progress in *T. cruzi* screening policies, both USPs and 100% coverage are yet to be achieved. Seroprevalence in BBs have decreased in endemic countries, likely due to a combination of vector control, increased USPs and voluntary donation, and improved diagnostic tests. To achieve the proposed WHO goals by 2025, USPs in TCs must become available in all endemic and non-endemic countries where migration from endemic countries is important.

## Data Availability

Data used in this paper is included in the manuscript and totally available.

## Contributions

JK, ZMC: Study design, formal analysis and first draft of the manuscript. JK, JL, ERM: abstract selection, and data extraction. MGB, PN, AD, MGB: revised and edited the manuscript.

## Acknowledgments

We thank the Imperial College London library service for the provision of essential full text papers non publicly available.

